# Integrating Mental Health and Psycho-Social Support (MHPSS) into infectious disease outbreak and epidemic response: an umbrella review and operational framework

**DOI:** 10.1101/2023.07.27.23293219

**Authors:** Muhammad Alkasaby, Sharad Philip, Zain Douba, Hanna Tu, Julian Eaton, Muftau Mohammed, Mohammad Yasir Essar, Manar Ahmed Kamal, Mehr Muhammad Adeel Riaz, Marianne Moussallem, William K Bosu, Ian Walker

## Abstract

**Introduction:** Infectious disease outbreaks have a substantial impact on people’s psychosocial well-being. Yet, mental health and psychosocial support (MHPSS) interventions are not systemically integrated into outbreak and epidemic response. Our review aims to synthesise evidence on the effectiveness of MHPSS interventions in outbreaks and propose a framework for systematically integrating MHPSS into outbreak response.

**Methods:** We conducted an umbrella review in accordance with the Joanna Briggs Institute (JBI) methodology for umbrella reviews.

**Results:** We identified 23 systematic literature reviews, 6 of which involved meta-analysis, and only 30% (n=7) were of high quality. Most of the available literature was produced during COVID-19 and focused on clinical case management and medical staff well-being, with scarce evidence on the well-being of other outbreak responders and MHPSS in other outbreak response pillars.

**Conclusion:** Despite the low quality of the majority of the existing evidence, MHPSS interventions have the potential to improve the psychological well-being of those affected by and those responding to outbreaks. They also can improve the outcomes of the outbreak response activities such as contact tracing, infection prevention and control, and clinical case management. Our proposed framework would facilitate integrating MHPSS into outbreak response and hence mitigate the mental health impact of outbreaks.

**Review registration:** PROSPERO CRD42022297138.

## Introduction

On 4^th^ May 2023, the World Health Organisation (WHO) announced that COVID-19 no longer constitutes a public health emergency of international concern (PHEIC)(1). This declaration came after more than three years, during which time the virus has accounted for more than 6.9 million deaths and nearly 760 million infections worldwide (2). Since it started, the COVID- 19 pandemic has had a considerable impact on people’s mental health (3). To reduce the number of infections and deaths, governments all over the world took precautionary measures such as travel and movement restrictions, and physical distancing. Paradoxically, physical distancing inadvertently exacerbated the impact of the COVID-19 pandemic on mental health (3–6).

The substantial effect of infectious disease outbreaks on mental health is not exclusive to the COVID-19 pandemic, as evidence linking mental health problems to earlier epidemics is widespread (7). Studies from Ebola Virus Disease (EVD) outbreaks document an increased incidence of mental health conditions such as depression, anxiety, stress, and post-traumatic stress disorder (PTSD) among the affected population, such as EVD survivors and their families, and healthcare workers, burial teams, etc. (8,9). Similarly, during the Middle Eastern Respiratory Syndrome (MERS) epidemic, a study from South Korea reported more than half of the survivors had at least one symptom of PTSD or depression and more than a quarter continued to have sleep difficulties even a year later (10). Healthcare providers were similarly affected (11). Following the severe acute respiratory syndrome (SARS) outbreak in 2002-03, a meta-analysis revealed that 28%, 20%, and 19% of the survivors were affected by clinical PTSD, depressive and anxiety disorders, respectively (10). Thus, there is clear evidence that mental health problems occur much more frequently amongst both survivors and healthcare workers during and after such epidemics compared with background rates in the population.

However, it must be noted that none of these epidemics became a pandemic and disrupted life as much as COVID-19. In 2020, there was an estimated increase of 25% (more than 100 million cases) in the global prevalence of both major depressive and anxiety disorders due to COVID-19 (12). The pandemic has also had a disruptive impact on mental health services worldwide. In a WHO survey, over 90% of 130 member states reported significant disruptions to their mental health services during the COVID-19 crisis (13). This ranged from difficulties in maintaining community supports, to medical service disruption and repurposing of mental health beds to COVID-19 wards. These profound impacts prompted the United Nations (UN), WHO, and other institutions providing normative guidance to recommend the inclusion of mental health and psychosocial support (MHPSS) in the COVID-19 response (14,15).

Despite this guidance, MHPSS was not systemically integrated into the COVID-19 response in many countries (16). In the WHO survey, although 89% of countries reported that MHPSS was part of their national COVID-19 response plans, only 17% of them ensured that full additional funding was available for MHPSS activities, demonstrating a gap between planning, funding, and implementation. In 28 countries across Africa, the degree of implementation of MHPSS activities recommended by the Inter-Agency Standing Committee (IASC) was less than 50% in most countries (16,17).

The COVID-19 pandemic has prompted many literature reviews on MHPSS interventions during infectious disease outbreaks. However, we are not aware of any published umbrella review of the evidence from these systematic reviews. Moreover, the existing literature rarely maps on to the coordination structure of outbreak response (pillars of the Incident management system) which is different to the structure used in humanitarian settings (the cluster approach), most common to MHPSS studies. We therefore aimed to (1) synthesise findings from systematic reviews and meta-analyses on the effectiveness of MHPSS interventions in the context of infectious disease outbreaks, (2) identify the different MHPSS interventions in relation to outbreak response pillars, and (3) propose a framework for systematically integrating MHPSS into infectious disease outbreak response.

### Review Questions

We had two main review questions:

- What are the MHPSS interventions that are deemed effective and feasible in the context of infectious disease outbreaks?
- How can these MHPSS interventions be integrated into the recognised outbreak response pillars?

### Inclusion Criteria

#### Types of participants

Review articles in which the study participants included people of all age groups and professions affected by infectious disease outbreaks, whether infected or not. The participants included (but was not limited to) children, adults, health care providers, volunteers, people with existing mental illness, people with disability, infectious disease patients (cases) and their carers/household/family, as well as the wider community.

#### Interventions

MHPSS interventions included any type of intervention that aims to protect or promote psycho-social well-being and/or prevent or treat mental disorders (18) and mitigate the effect of infectious disease outbreaks on mental health.

#### Context

The current umbrella review covered interventions implemented in the context of infectious disease outbreaks of different scales (e.g. EVD and SARS) up to a global pandemic such as COVID-19.

#### Outcomes

We included Systematic literature reviews that reported mental health-related outcomes. The main outcomes included (but were not limited to) stress, depression, anxiety, and post-traumatic stress disorder (PTSD). Other outcomes were resilience, coping, and quality of life. Measures of these outcomes included self-reports of mental health and well-being, use of mental health and psychological screening instruments, and psychiatric diagnostic interviews. We also included implementation outcomes related to the acceptability and feasibility of interventions (if available).

#### Types of studies

We limited our selection to peer-reviewed systematic literature reviews and meta-analyses. Reviews that did not report a systematic method of literature search and selection were excluded. Only reviews that exclusively or partially covered interventional studies were included. We included only reviews that had a full-text in English.

## Materials and Methods

Umbrella reviews synthesise review-level evidence from the published literature and are increasingly common in public health research to summarise evidence for a given topic. This umbrella review was conducted in accordance with the Joanna Briggs Institute (JBI) methodology for umbrella reviews (19). The review protocol was registered with PROSPERO (PROSPERO CRD42022297138).

### Search Strategy

We identified keywords for mental health interventions, outbreaks, and mental health-related outcomes and conducted a preliminary search on PubMed to test our strategy. After that, we searched the following electronic databases: PubMed, Scopus, Web of Science, CINAHL, Cochrane, Embase, Epistemonikos, Global Health, MEDLINE, and PsycInfo for relevant studies published until January 2022. Furthermore, We searched the reference lists of the included review for potentially relevant articles. We did not use time or language limits in our search strategy (though were unable to analyse if full text was not available in English). The detailed search strategy for each database and the number of studies retrieved can be found in the Supplemental Files (S1).

#### Study screening and selection

After removing duplicate citations using the EndNote reference management software, reviewers, working in pairs, independently screened the titles and abstracts of the included studies guided by the eligibility criteria. A senior reviewer (IW) was consulted to resolve any conflicts in the screening results and made the final decisions about retrieval for full-text review. The pairs of reviewers continued to screen the retrieved full-text articles independently, and the senior reviewer consulted to resolve any conflict.

### Assessment of methodological quality/critical appraisal

To assess the methodological quality of the included reviews, we used the JBI critical appraisal instrument for Systematic Reviews and Research Syntheses (19). We agreed upon the following cut-off scores based on fulfilling the 11 items of the instrument: less than 6 (low quality), 6-8 (moderate quality), and over 8 (high quality). Two reviewers independently assessed the quality of the included study and a third reviewer resolved any conflicts.

### Data collection

We developed an data extraction tool using an Excel template. After discussing it with all reviewers, we piloted it before starting the actual extraction of data to maximize consistency between all reviewers and to ensure that all relevant information was extracted. Pairs of researchers extracted the data from the included studies and resolved any discrepancies by consensus. The extracted data included first named authors, year published, objectives of the included review, participants’ characteristics, context or geographic setting, date of database searching, type of study, countries of origin of the included primary studies, instrument used to appraise the primary studies and their quality rating, the outcomes reported that were relevant, and the method of synthesis/analysis employed to synthesize the evidence.

### Data Summary

As the data in the included reviews were too heterogenous to synthesize quantitatively through meta-analysis, we used narrative synthesis to summarise the findings. To identify effective MHPSS interventions in the context of outbreaks, we extracted findings - reported in the included reviews - from RCTs of MHPSS interventions targeting different population groups affected by an outbreak. We then conducted a qualitative synthesis for the included reviews grouping interventions according to the outbreak response pillars and the target population. We sought to develop a model of MHPSS interventions from the findings that could be applied to outbreak response pillars.

### Deviations from the review protocol

Initially, we planned to group interventions according to target population, but after consulting subject matter experts, and given our goal, which is integrating MHPSS into outbreak response, we decided to group interventions according to outbreak response pillars.

## Results

### Study inclusion

We initially identified 1,883 records, out of which 106 potentially met our inclusion criteria for full-text screening (Figure 1). Of the latter, 23 records met the inclusion criteria and were finally included in the qualitative synthesis.

**Figure 1:**
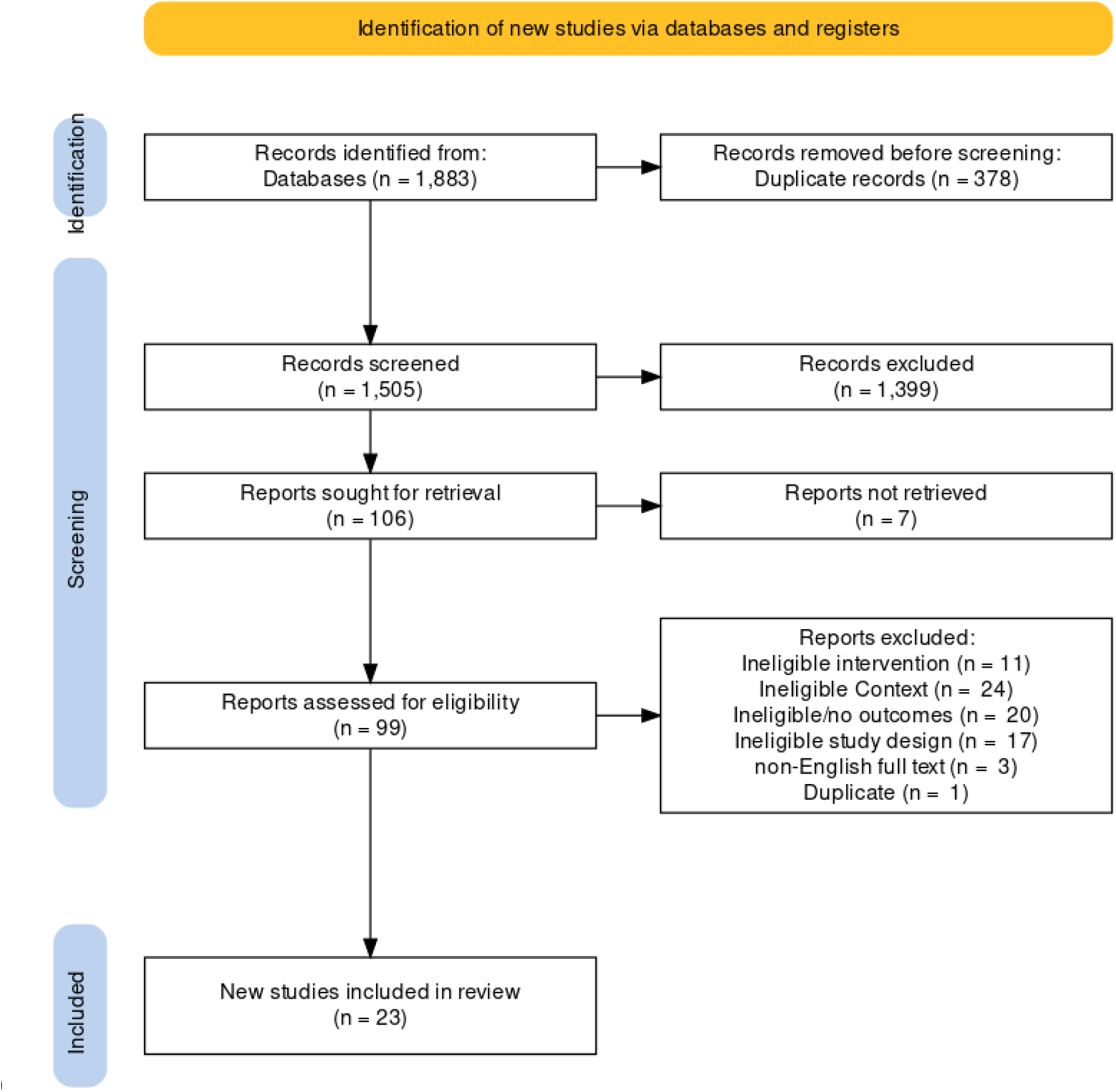
PRISMA Flow Diagram.

### Characteristics of the included study

All the included studies were systematic literature reviews, with six of them involving meta-analysis (Table 1). Only three reviews included RCTs exclusively (20–22). The included studies covered 47 countries; 27 of them (57%) were high-income countries (HIC), while 11% were low-income countries (LIC) (Figure 2).

**Figure 2:**
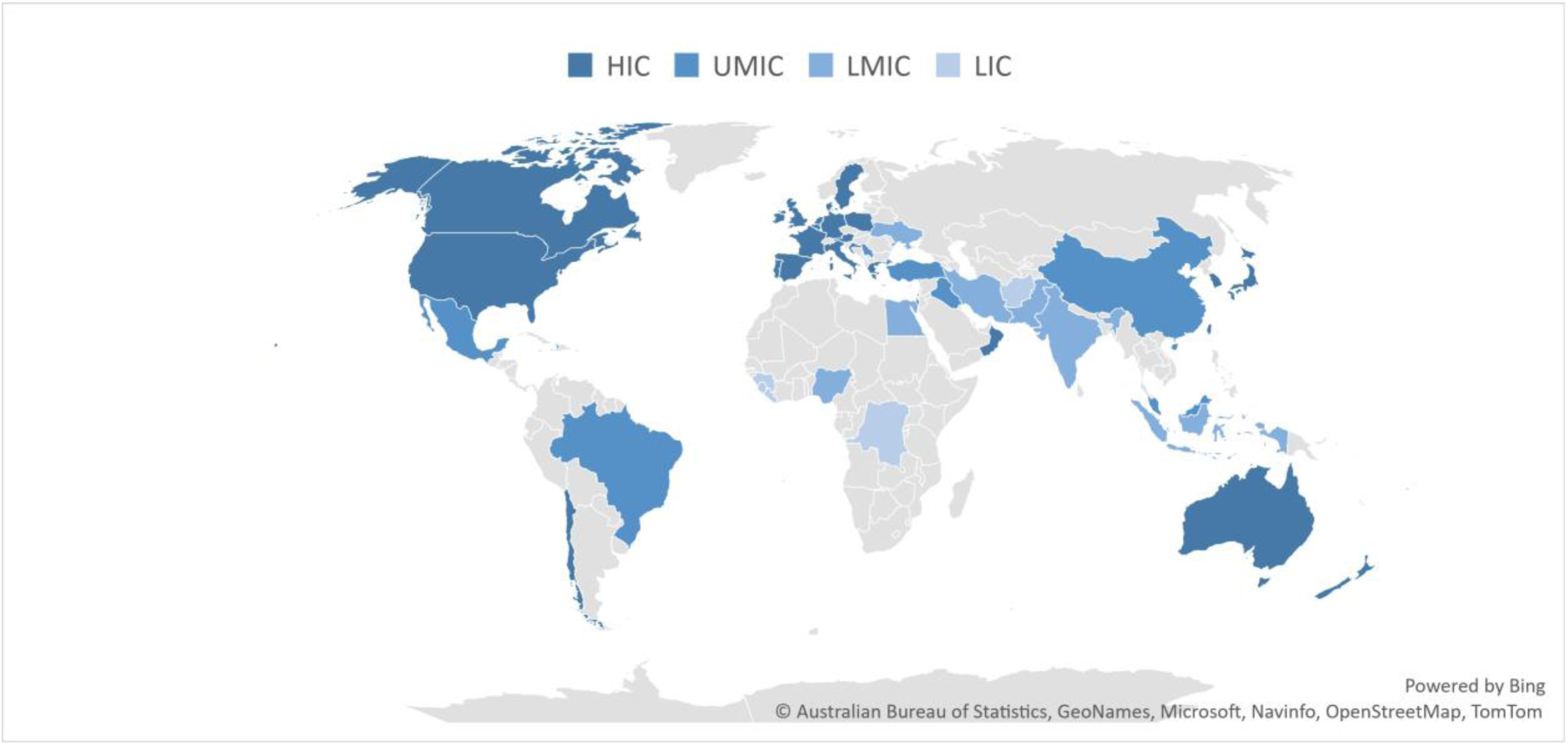
Countries covered by the included reviews according to the income category.

**Table 1:** Summary of the Included Reviews.

| Author, Year | Type of Review | Objective | Setting | Conclusion and Recommendations | Overall Quality Assessment |
| --- | --- | --- | --- | --- | --- |
| (Bertuzzi *et al.*, 2021) (28) | Systematic Review | To provide a summary of the evidence for the available psychological support interventions and strategies for HCWs and informal caregivers during COVID-19. | COVID-19 | Digital interventions were feasible and efficient in providing psychological support. Usability and digital literacy should be considered when designing digital interventions | High Quality |
| (Bursky *et al.*, 2021) (27) | Systematic Review | To summarize the findings and draw research-based recommendations for the application of meditation interventions for prisoners and individuals in quarantine or lockdown during the current pandemic. | COVID-19 and other settings that resembles quarantine and isolation in outbreaks | Meditation can be utilized during quarantine to mitigate negative feelings of loneliness, fear, and worry, improve psychopathological symptoms such as depression and anxiety, bolster physiological functioning, and increase overall well-being, allowing for a potentially positive transformative quarantine experience. | Moderate Quality |
| (Cénat, Felix, *et al.*, 2020) (9) | Systematic Review | To describe the implementation of MHPSS programs following the past EVD outbreaks | Ebola | There is a need to systematically document and evaluate the implemented MHPSS programs. Research could help understand the complex relationship between the perception of a threat, the associated psychological distress and the adoption of preventive behaviours in specific social and cultural environments. | Low Quality |
| (Damiano *et al.*, 2021) (29) | Systematic Review & Meta-analysis | To review the most common mental health strategies aimed at alleviating and/or preventing mental health problems in individuals during the coronavirus disease 2019 (COVID-19) and other coronavirus pandemics. | COVID-19, SARS, and MERS | The meta-analysis (of 3 RCTs) revealed that the interventions promoted better overall mental health outcomes as compared to control groups. The review identified a large body of expert recommendations, however, most articles had a low level of evidence | Moderate Quality |
| (Davison *et al.*, 2021) (30) | Scoping Review | To describe the effective health promotion, primary prevention, screening, and treatment interventions to enhance mental | COVID-19 | Only one pilot study targeted the mental health of those with chronic health conditions. Patient navigator programs, | Moderate Quality |
|  |  | health outcomes and reduce the risk of substance use for populations with chronic physical health conditions who are at risk of contracting COVID-19 and having severe symptoms |  | group online medical visits, peer support, and social prescribing may support those with complex needs. Future policies need to address digital health access inequities and the implementation of multi-integrated health and social care. |  |
| (Ding *et al.*, 2021) (20) | Systematic Review & Meta-analysis | To evaluate the effects of non-drug interventions on anxiety, depression and sleep in patients with COVID-19 | COVID-19 | This meta-analysis found that non-drug interventions can reduce the anxiety and depression scores of COVID-19 patients. | High Quality |
| (Doherty *et al.*, 2021) (21) | Systematic Review & Meta-analysis | To assess the effectiveness of psychological interventions in the general population and healthcare workers exposed to infectious disease outbreaks | COVID-19 and SARS | Meta-analysis showed that psychological interventions had a statistically significant benefit in managing depression and depression. Psychological interventions are needed for those vulnerable to the mental health consequences of outbreaks | High Quality |
| (Gómez *et al.*, 2021) (23) | Rapid systematic review | To review evidence on mental health interventions for children exposed to community crises or disasters | COVID-19, conflicts, and natural disasters | Involve different actors from the education sector in developing develop interdisciplinary and participative mental health strategies. Training teachers to implement interventions could increase the system capacity to reach and follow the children over a long time period. | Low quality |
| (Hooper *et al.*, 2021) (24) | Systematic Review | To identify and summaries recent early psychological intervention programs that were administered to prevent or minimize psychological harm in frontline responders | COVID-19 | PFA, EMDR, and trauma risk management showed effectiveness across at least two studies each with frontline workers. Organisations have a duty of care to support their staff and equip them with psychological skills to help them cope with the mental health impact of crises | Low Quality |
| (Kunzler *et al.*, 2021) (31) | Scoping Review | to identify and summarize the available literature on interventions that target the | Ebola, MERS, SARS, | Most interventions delivered focused on healthcare workers and crisis personnel and | Moderate Quality |
|  |  | distress of people in the face of highly contagious disease outbreaks | COVID-19, and Influenza | combined psychoeducation with training on coping strategies.<br>There is a need for pandemic preparedness interventions and interventions that target specific target groups (e.g., children) |  |
| (Meherali *et al.*, 2021) (32) | A Rapid Systematic Review | To advise public health and policymakers on strategies and interventions to improve mental health among children and adolescents during pandemics | Ebola, Equine Influenza, H1N1, and COVID-19 | Interventions such as art-based programs, support services, and clinician-led mental health and psychosocial services effectively decrease mental health issues among children and adolescents.<br>Age-appropriateness and channel of delivery should be considered | Moderate Quality |
| (Pollock *et al.*, 2020) (33) | Systematic Review | To assess the effects of interventions aimed at supporting the resilience and mental health of frontline health and social care professionals during and after a disease outbreak, epidemic or pandemic and identify barriers and facilitators that may impact on the implementation of these interventions | Ebola, SARS, MERS, COVID-19 | MHPSS interventions for HCWs should address organisational, social, personal, and psychological factors. PFA training can strengthen HCWs' capacity to provide psychosocial support in disasters and humanitarian crises. | High Quality |
| (Puyat *et al.*, 2020) (25) | A Rapid Systematic Review | To examine activities that can promote mental wellness during pandemics, quarantines, social isolation, or other stress-inducing Events. | COVID-19 and other settings that resembles quarantine and isolation in outbreaks | Physical activity such as exercise and yoga can improve mental well-being during outbreaks. Such interventions should be offered in conjunction with conventional mental health services when directed towards persons with pre-existing mental health conditions. | Moderate Quality |
| (Rivera-Torres *et al.*, 2021) (34) | Scoping Review | To map evidence on the types of leisure and recreation activities (LRA) older adults, 60 years and older, are engaged in for their | COVID-19 | Older adults benefited from both digital (such as social networks) and physical (such as walking outside, aerobics, gardening) LRA activities. Social connectedness was | Low Quality |
|  |  | mental health during the COVID-19 pandemic. |  | beneficial, especially for those at higher risk for social isolation and loneliness. |  |
| (Serrano-Ripoll *et al.*, 2020) (35) | Systematic Review & Meta-analysis | To examine the impact of health emergencies caused by a viral pandemic or epidemic outbreak on HCWs' mental health; ii) to identify factors associated with worse impact; and iii) to assess the effectiveness of interventions to reduce such impact. | COVID-19, SARS, MERS, H1N1 Influenza, H7N9 Influenza, and Ebola | Four studies reported interventions for frontline HCW: two educational interventions increased confidence in pandemic self-efficacy and in interpersonal problems solving (very low certainty), whereas one multifaceted intervention improved anxiety, depression, and sleep quality (very low certainty). | High Quality |
| (Shatri *et al.*, 2021) (36) | Clinical Review | To conduct a clinical review of the role of online psychotherapy in patients with COVID-19. | COVID-19 | Providing mental health support, especially via telehealth, is likely to help patients maintain psychological well-being and better cope with health impacts of COVID-19 | Low quality |
| (Soklaridis *et al.*, 2020) (37) | A Rapid Systematic Review | To Report on implementation, evaluation, and outcomes regarding mental health interventions during medical pandemics within the last two decades. | MERS, SARS, Influenza Pandemics, Ebola, And COVID-19 | Attention needs to be paid to cultural considerations when designing and implementing mental health interventions and training. Training non-specialists empowers communities to deliver mental health interventions. | Moderate Quality |
| (Strudwick *et al.*, 2021) (38) | Rapid systematic review | What digital interventions could be used to support the mental health of the Canadian general population during the COVID-19 pandemic? | COVID-19 and other disasters | Indigenous peoples should be included at every stage of the intervention development. Ensure cultural relevance, equity and access when developing digital interventions | Moderate Quality |
| (Sun *et al.*, 2021) (22) | Systematic Review & Meta-analysis | To evaluate the effect of psychological interventions on HCWs with PTSD due to their exposure to life-threatening pandemics | Life-Threatening Pandemics such as COVID-19 | The most effective and feasible treatment option for HCWs with PTSD is still unclear, but CBT and MBX have displayed the most significant effects based on currently limited evidence. | Moderate Quality |
| (Williams *et al.*, 2021) (26) | Rapid systematic review | To evaluate the current evidence base for interventions deemed compatible with shielding/social distancing measures | Settings that resemble quarantine | A combination of educational and psychological approaches that target the root cause of one's loneliness, in addition to social facilitation initiatives, to create and maintain | Moderate Quality |
|  |  |  | and isolation in COVID-19 | relationships, represent the best opportunities to improve loneliness and decrease the impact of public health measures during outbreaks |  |
| (Yang *et al.*, 2021) (39) | Systematic Review & Networking Meta-analysis | To compare the different psychological interventions and identify the most effective way to treat the psychological manifestations in people affected by COVID-19 | COVID-19 | A comprehensive analysis of the results indicated that supportive therapy was the most commonly used therapy and showed a better performance in anxiety and depression measurement scales | Moderate Quality |
| (Yue *et al.*, 2020) (40) | A Rapid Systematic Review | To synthesize the data on mental health services and interventions for the infectious disease epidemics and to enhance knowledge and improve the quality and effectiveness of the mental health response to COVID-19 and future infectious disease epidemics. | COVID-19, SARS, EVD, and MERS | Group-based CBT, PFA, community-based psychosocial arts program, and other culturally adapted interventions are effective against the mental health impacts of outbreaks. Mental health strategies integrated into public health emergency response can enhance response capacity for outbreaks. | Moderate Quality |
| (Zaçe *et al.*, 2021) (41) | Systematic review | To describe interventions that have been implemented to tackle mental health issues in HCWs during infectious disease outbreaks and measure the efficacy of these interventions | SARS, Influenza A, H1N1, Ebola, COVID-19 | Facilitators and barriers to the implementation of interventions should be identified and considered in the outbreak planning process. Interventions for HCWs should not only focus on the individual-level factors, but should also target organisational and system-related factors | Moderate Quality |
| CBT= Cognitive Behavioural therapy EMDR= Eye Movement Desensitisation and Reprocessing; EVD= Ebola Virus Disease; HCWs= Healthcare workers; LRA= Leisure and Recreation Activities; MHPSS; Mental Health and Psychosocial Support; MBX = Mindfulness-Based Stretching and Deep Breathing Exercise; PFA= Psychological First Aid; PTSD= Post-Traumatic Stress Disorder; RCTs= Randomized Controlled Trials; SARS= Severe Acute Respiratory Syndrome |  |  |  |  |  |

All the included reviews addressed exclusively or partially MHPSS intervention in the context of outbreaks. Some of the reviews included interventions from other contexts that have the potential to be effective in the context of outbreaks (such as incarceration, which resembles quarantine) (23–27).

### Methodological quality appraisal

Based on our quality appraisal criteria, we judged that 21.7% of the included reviews (n=5) were of high quality, 56.5% were of moderate quality (n=13), and 21.7% were of low quality (n=5). The overall quality appraisal result of each study is shown in Table 1. The quality appraisal tool and detailed quality appraisal results for the included studies can be found in the Supplemental Files (S2). Most of the primary studies included in the reviews were not randomised controlled trials (RCTs) and included a wide range of study designs, such as quasi-experimental and observational studies. The quality of evidence from the included primary studies was generally low, as reported by the included reviews’ authors. Limitations included small sample sizes, lack of randomisation and blinding, and high loss of follow-up.

### Findings in relation to infectious disease outbreak response pillars

The included reviews reported the results of 26 RCTs (Table 2), in addition to other types of experimental and non-experimental studies. To facilitate the integration of MHPSS into outbreak response, we present findings of this umbrella review in relation to the infectious disease outbreak response pillars.

**Table 2.**
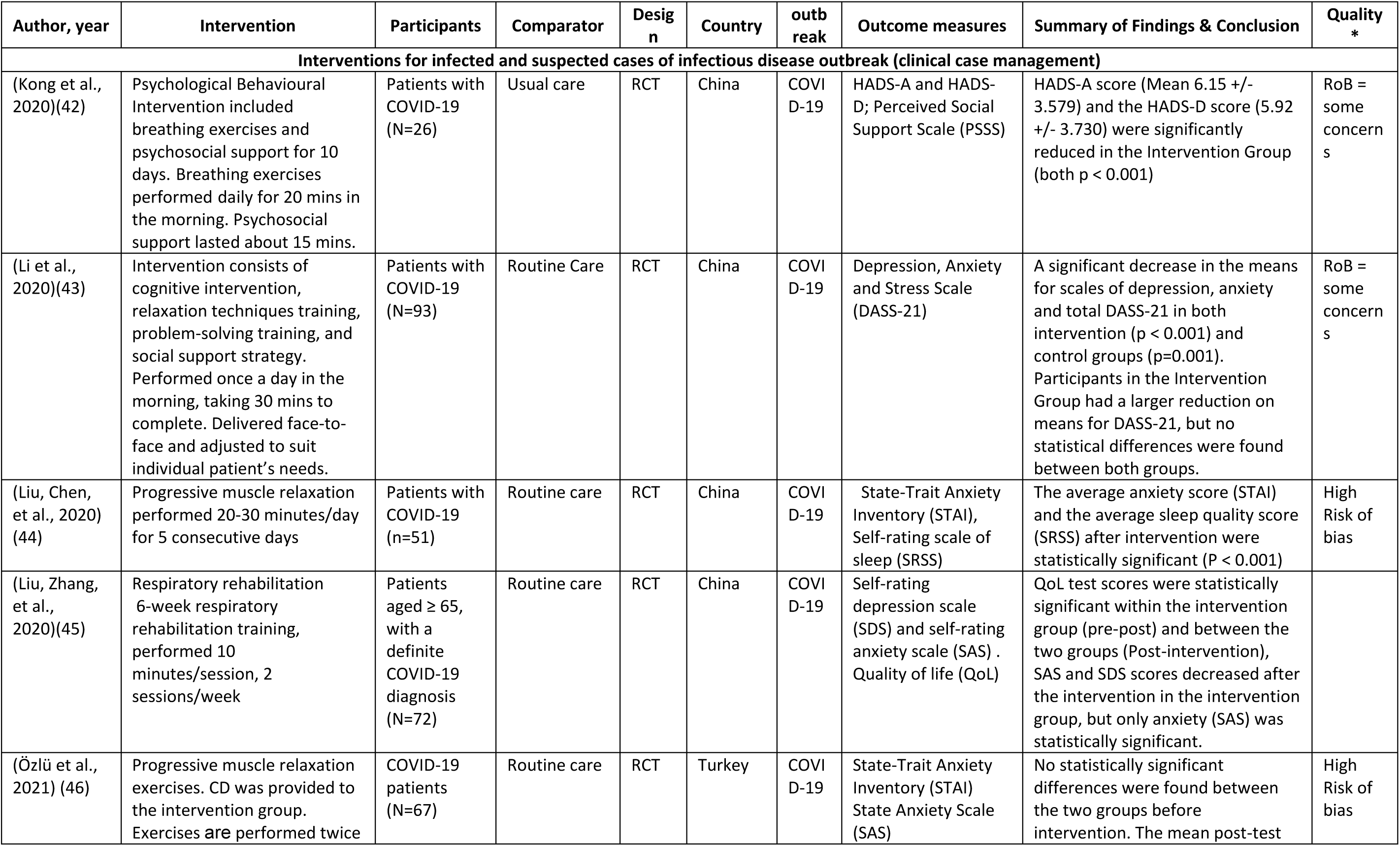
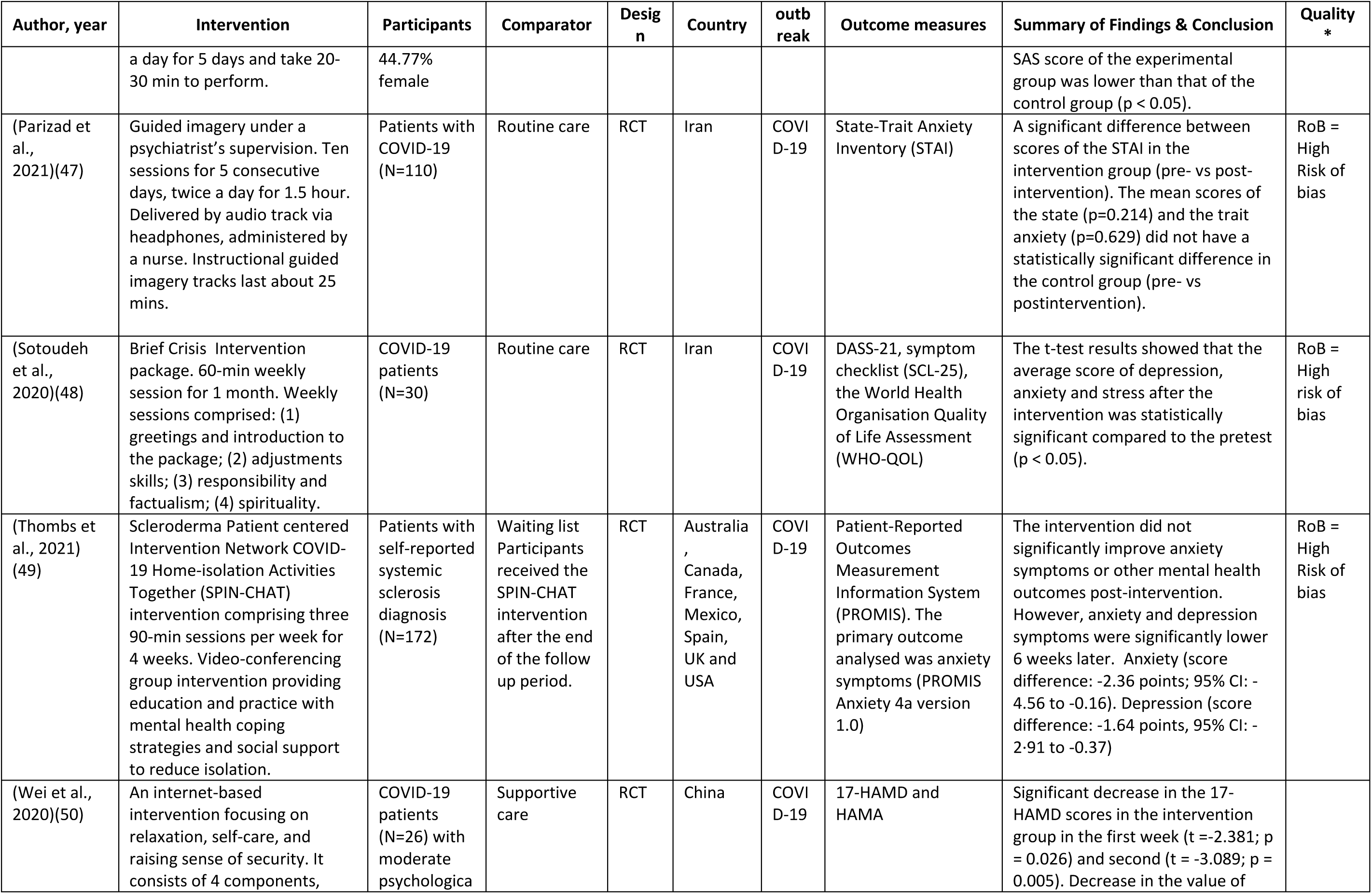
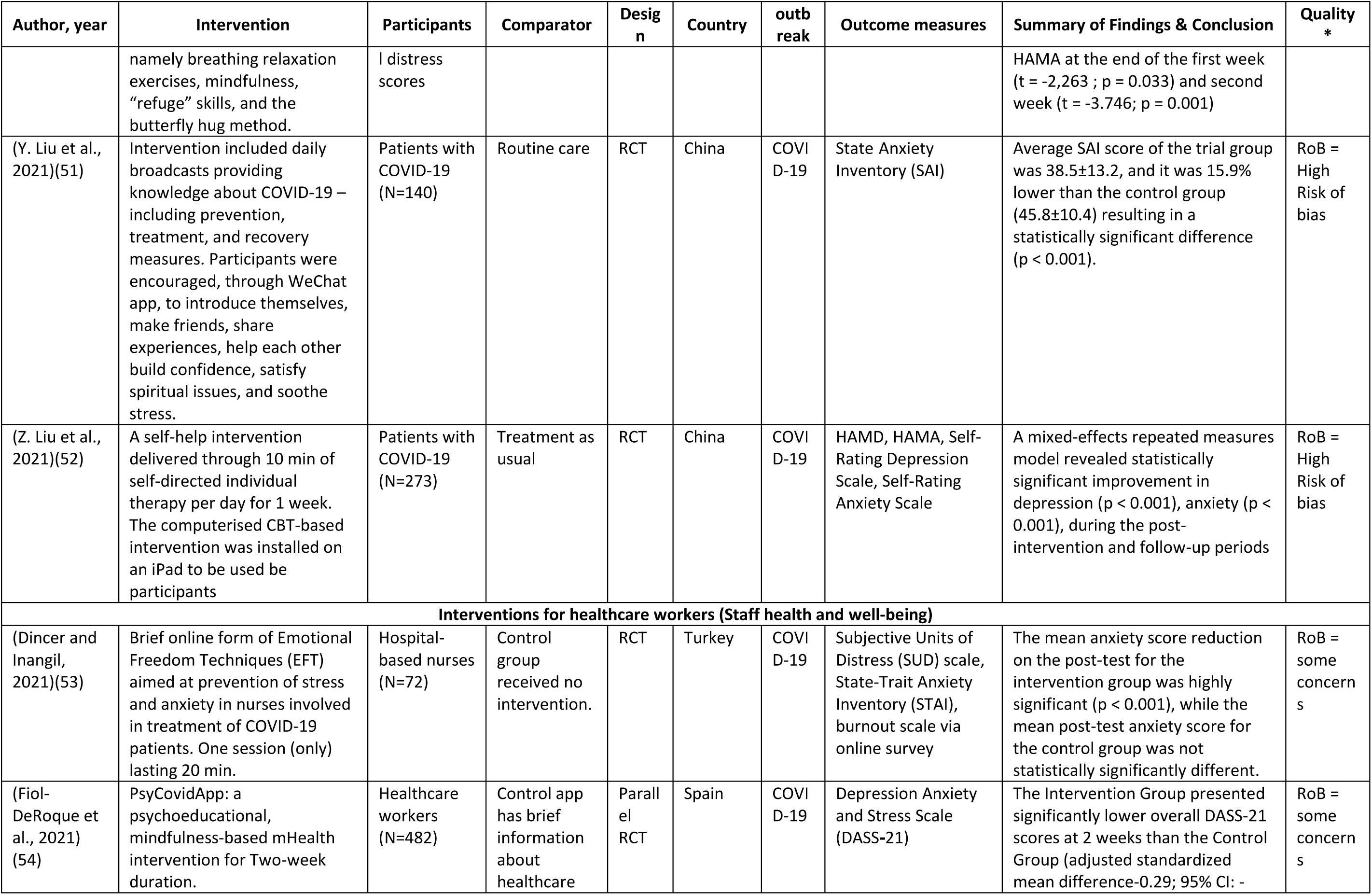
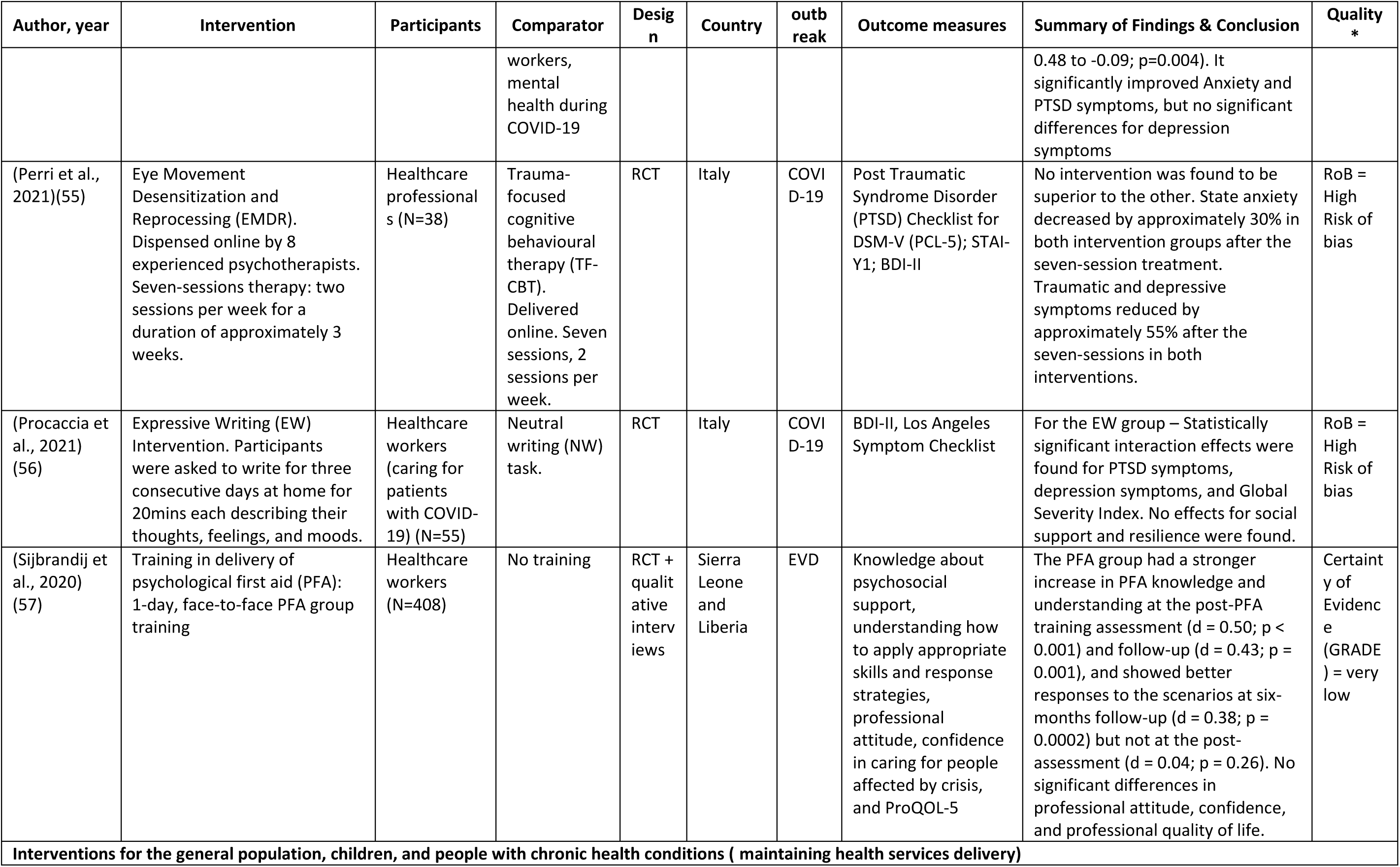
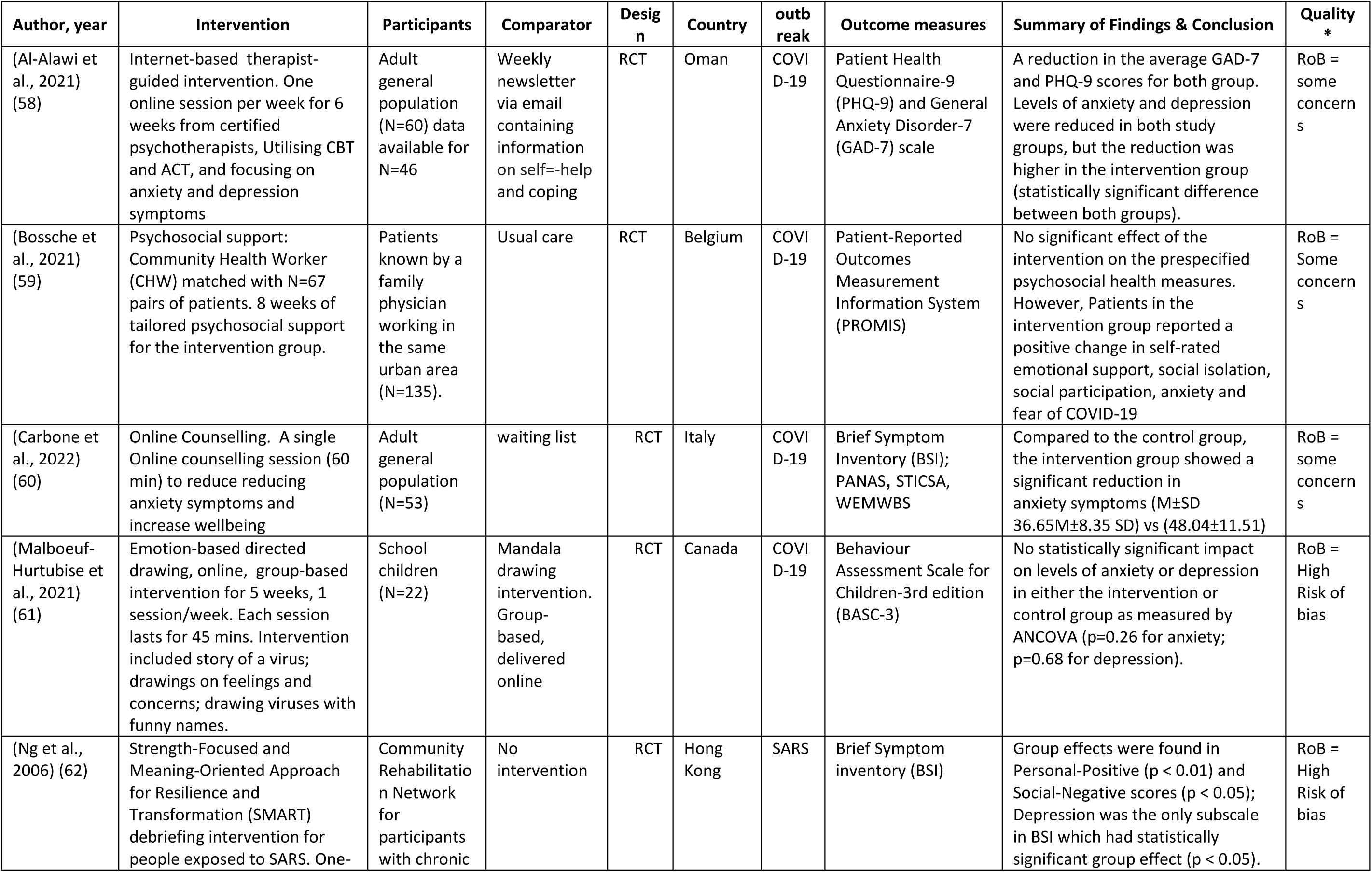
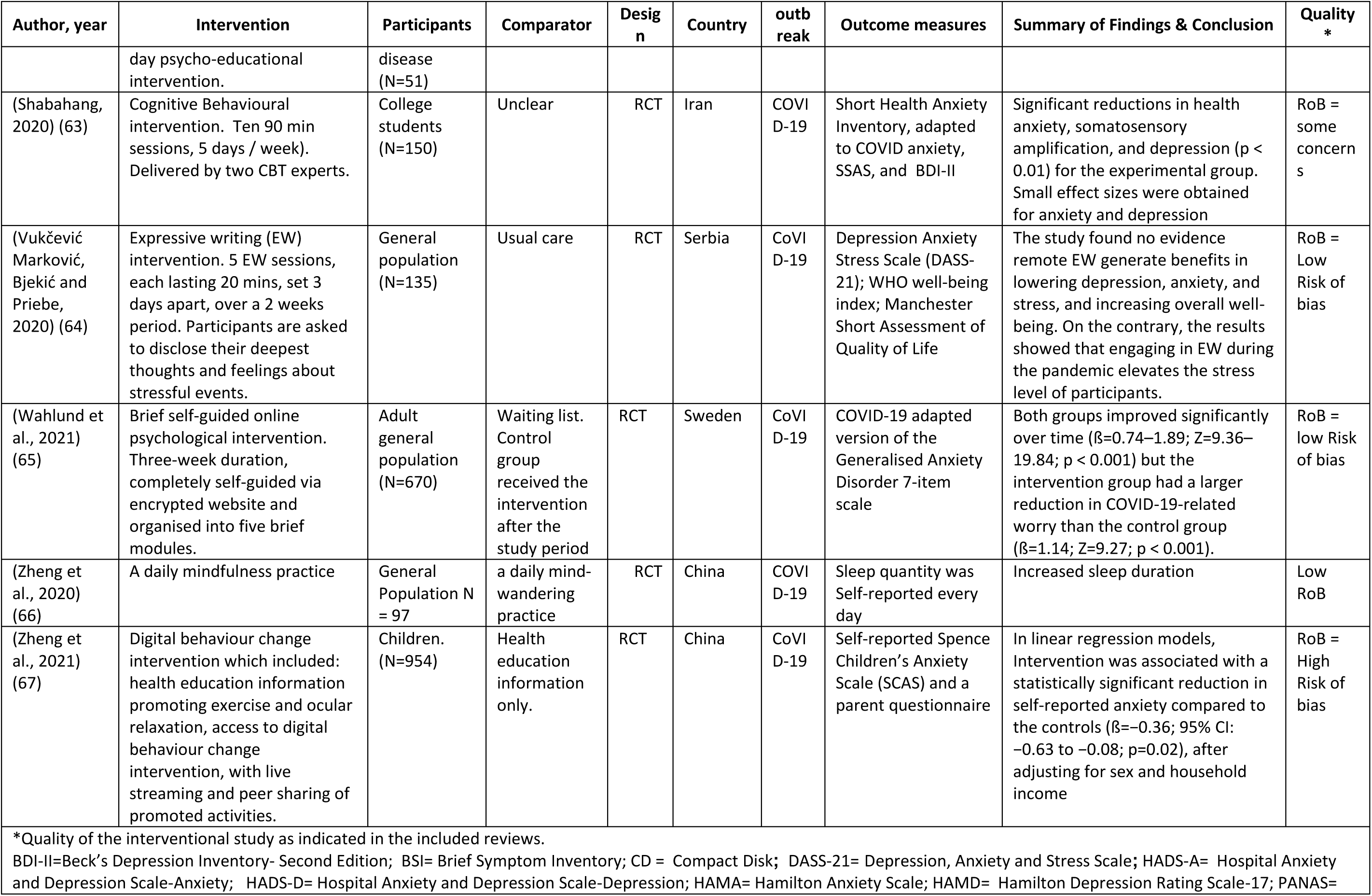
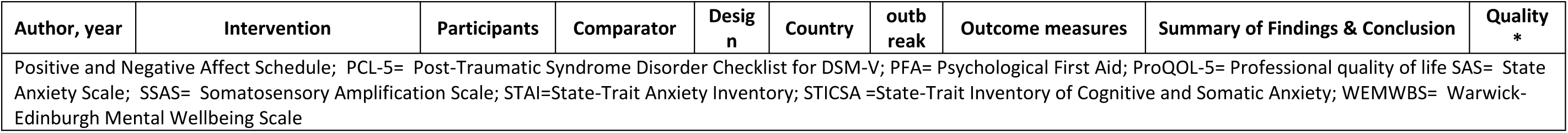
Summary of RCTs reporting MHPSS interventions in the context of infectious disease outbreaks.

#### 1. Multisector and Partner coordination

Partner coordination is essential to ensure the appropriate coverage and quality of services for the affected population, especially vulnerable groups. It also ensures the appropriate use of limited resources available, avoids duplication (68) and coordinates the response across multiple sectors (17,69).

Education was one of the sectors identified in our review that implemented MHPSS activities during outbreak response. One review reported interventions for children returning to school after crisis (23). Out of the 18 included interventions, 12 (66.7%) were school-based and only one intervention was conducted in an outbreak context. Evidence from other health crises was used to inform recovery and back-to-school strategies after the COVID-19 pandemic (23). Most of the interventions were school-based, used cognitive-behavioural therapy (CBT) techniques, and were delivered by teachers, therapists, and clinicians (23). Another review reported an example of multisectoral collaboration during Ebola outbreak in West Africa, where mental health clinicians and law enforcement forces worked collaboratively to improve the outbreak response (40). Healthcare workers and police forces were trained on Psychological First Aid (PFA) and nonviolent de-escalation techniques that could be used for agitated patients, especially those in Ebola Treatment Units (ETUs), and improve their engagement with the public (40). This coordination also facilitated referrals to mental health services for those needing further intervention (40,70). Overall, the health sector generated most of the MHPSS evidence in outbreak response, with little evidence from other sectors on cross-sectoral activities.

#### 2. Staff health and well-being

Six (26.1%) reviews addressed frontline staff well-being (22,24,28,33,35,41). While most of the reviews on frontline workers focussed on health care workers’ well-being, one review included other frontline (non-health) responders (24). In a Cochrane review that investigated MHPSS interventions for health and social care professionals during outbreaks, one interventional study that assessed the impact of PFA training on healthcare workers was included. According to the authors, the included study provided a very low certainty of evidence on the impact of workplace interventions on burnout among healthcare workers (33). The Cochrane review found no interventions for social care workers’ well-being during outbreaks. Authors suggested considering evidence from other emergencies due to the lack of available evidence from studies conducted in the context of outbreaks (33).

Training frontline workers in PFA has a potential benefit not only for those who will receive PFA but also for those who are trained to provide it. In one included review, PFA was reported to improve the positive mental health outcomes of health care workers and other first responders, such as resilience, coping, self-efficacy, perceived social support, and reduced perceived self-stigma (24). It also improved the trainees’ knowledge, understanding, and skills regarding providing support to affected individuals (24).

PFA training was not limited to medical staff only, but it also included others involved in the outbreak response. In the Ebola outbreak in west Africa, community and faith leaders, police, contact tracers, volunteers, Ebola survivors, social mobilization agents, and food providers to quarantined households were trained in PFA (71).

Peer-support was one of the frequently recommended approaches for staff well-being (72–75). The “buddy system” is a peer support model where two people who work together are able to monitor and support each other. In addition to the psychosocial benefits for the workers, peer support can have a technical benefit where less experienced personnel can learn quickly and closely from experienced colleagues (72).

In terms of disorder-specific interventions, a review (22) investigating MHPSS interventions for health personnel working in hospitals and emergency services who suffer from PTSD symptoms, reported significant improvement in the PTSD symptoms following exposure-based CBT and Mindfulness-Based Stretching and Deep Breathing Exercises (76,77). However, the level of evidence reported in that review was evaluated as low.

#### 3. Infection prevention and control (IPC) measures

IPC involves the measures taken to prevent the transmission of an infectious agent. This includes the use of personal protective equipment (PPE), hand hygiene, and environmental cleaning. It also includes safe burial practices in some outbreaks, such as Ebola. Included reviews reported some adaptations to MHPSS activities to be delivered safely during outbreaks. For instance, in addition to following IPC protocols, mental health services postponed unnecessary outpatient visits and replaced them with check-ins via phones; group activities were suspended or delivered remotely (e.g., using Zoom); family visits to in-patients were restricted (40,78). Since staff might be the only people in contact with patients in mental health facilities, electronic tracking of staff movement was applied in one of the studies to facilitate contact tracing (40,79). Another study reported a home hospitalisation model where people with severe mental illness who need admission were taken care of at home by a mental health team (40,80).

The use of remote interventions has witnessed a remarkable increase during COVID-19, in line with infection prevention and control measures. Many psychiatric and counselling services shifted to digital provision. One review summarised that providing psychotherapy and counselling services online can reduce anxiety and depression, especially in patients experiencing mild to moderate symptoms (36). Remote interventions included in reviews varied regarding length, delivery agent, and content. However, a 60-minute single online counselling session delivered by a clinical psychologist and focused on psycho-education, anxiety management strategies, and empathic listening was found to be effective in reducing anxiety and negative affect (60).

In some infectious disease outbreaks, such as Ebola, where deceased patients are highly infectious, MHPSS interventions included providing support to families of the deceased and ensuring safe and dignified burials (71).

#### 4. Case management

Case management focuses on providing care and treatment for infected individuals and suspected cases who have been in contact with infected individuals. Integrating MHPSS into case management includes providing MHPSS to infected individuals and their contacts, especially those who are socially isolated. This can be provided remotely or in person (for those in healthcare facilities) after taking the necessary IPC measures to limit transmission.

Eleven RCTs of MHPSS interventions for infected patients were identified and extracted from the included reviews, all of which were conducted during the COVID-19 pandemic. A systematic review and meta-analysis that included five RCTs conducted in China with a total of 768 COVID-19 patients found that non-pharmacological interventions can improve symptoms of depression and anxiety in COVID-19 patients (20). Interventions included progressive muscle relaxation, respiratory rehabilitation, life intervention, nursing with traditional Chinese medicine and internet-based integrated interventions (20). Progressive muscle relaxation exercises effectively reduced anxiety and improved sleep quality in a sample of patients with COVID-19 in Turkey (46). Other interventions included CBT, guided imagery, and self-help interventions (21). While some interventions were provided in person, others used technology such as recordings or tablets.

Training for staff working with infected people and their contacts is an essential component of MHPSS interventions for case management, and it was reported frequently in the included reviews (70,71,81). During the Ebola outbreak in West Africa, those working in ETUs, Ebola survivors, police, and Psychosocial Support Teams received MHPSS training, including PFA and self-care (70,71,81).

#### 5. Maintaining essential health services

Outbreaks cause a significant increase in the prevalence of mental health conditions and disrupt mental health services (3,12). This necessitates integrating MHPSS into essential health services to ensure that people with existing mental health conditions, those with other co-morbid chronic conditions, and those who newly develop a mental health condition receive appropriate care. However, despite the high prevalence of mental health problems among patients with chronic health conditions, one review addressing the mental health of this population group during COVID-19 found only one intervention study targeting COVID- 19 patients, among whom 35% had chronic health conditions (30). Another review reported a video-conferencing group intervention for scleroderma patients during COVID-19 (49), and a group debriefing intervention for people with chronic diseases affected by SARS (62). Another important consideration to deliver MHPSS services within essential services during outbreaks is that services should be adapted to the existing IPC measures to limit the risk of transmission and ensure the safety of patients and services providers (see section under the IPC pillar above).

#### 6. Risk communication and Community Engagement (RCCE)

The RCCE pillar in an outbreak involves the effective communication of information about the outbreak to the public, as well as engagement with the community to mitigate the impact of outbreaks on their well-being and livelihood. During outbreaks, public health authorities might impose some measures to control the infection. Those measures can be associated with negative mental health impacts. Integrating MHPSS into RCCE includes communicating the risk of outbreaks on people’s psychosocial well-being and how to mitigate this risk (e.g., stress management and coping mechanisms). In general, there was a lack of literature on RCCE activities targeting vulnerable groups such as children and adolescents, older persons, caregivers, and people with psychosocial and physical disabilities.

In one of the primary studies, healthcare workers were trained to increase their preparedness for the H1N1 influenza pandemic (31,35,41,82). Training included information about the infectious agent, normal stress response, and coping mechanisms (82). The percentage of healthcare workers who felt confident about dealing with the pandemic increased from 35 % to 76% (82). Another review included studies that used text messages to support populations affected by the COVID-19 pandemic (40). Subscribers received free daily supportive messages for three months. Messages were prepared by a team of mental health professionals and service users (83,84). Participants in the intervention group receiving these messages had lower prevalence of stress and depression compared to the control group (84).

Given the impact of quarantine and lockdown on well-being, RCCE activities should address the psychosocial impact and provide evidence-based recommendations to mitigate the negative impact of these measures. Several reviews identified interventions that can be performed, in adherence to the IPC measures, to mitigate the negative impact of lockdown and isolation (25–27). In their review, Williams *et al.* (2021) investigated interventions for loneliness and social isolation that can be applied in the context of the COVID-19 lockdown. Mindfulness-based interventions, meditation, and laughter therapy were among the most effective interventions for loneliness and social isolation. Puyat *et al.* (2020) explored similar contexts to lockdown, such as prisons and summarised that activities such as exercise and yoga have beneficial effects on the mental health of prisoners.

Another review (27) investigated the effectiveness of meditation and mindfulness-based interventions for prisoners and people affected by lockdown and summarised that meditation and mindfulness-based interventions were effective in reducing symptoms related to stress and trauma among prisoners. Such activities could be performed at home easily with a little guidance and could be suggested as ways to reduce the negative impact of lockdown and other infection control measures.

## Discussion

This umbrella review identified limited evidence on MHPSS interventions in the context of outbreaks, epidemics, and pandemics. Many interventions were identified, but few were considered to demonstrate strong evidence. Despite experiencing several epidemics and pandemics in the last two decades, most of the available literature was produced during the recent COVID-19 pandemic. Moreover, the majority of studies were carried out only in healthcare settings which limits the scope of our findings. Research on MHPSS in outbreaks often focuses on healthcare staff well-being and providing psychological support for infected patients as part of clinical case management. In the sections below, we discuss the review findings according to outbreak response pillars and propose a model for integrating MHPSS into outbreak response in light of the exciting literature, guidelines and our technical expertise.

### MHPSS activities in outbreaks response

Despite the significant impact of public health emergencies on those involved in the response, the majority of the existing evidence we found focused on medical staff’s well-being, with little attention to others involved in the response (e.g., police, social services). The peer support approaches (e.g., buddy system) for staff well-being, which have important psychological, social, and technical benefits, are frequently mentioned in the literature (72–75). The buddy system was also used by the WHO during the Ebola outbreak in West Africa as a part of a holistic approach to ensure the occupational health and safety of emergency response personnel (85,86).

Healthcare workers are at high risk of contracting and transmitting the infection, which further exposes them to stigma and social isolation. Therefore, WHO recommends that public awareness campaigns address outbreak-related stigma and encourage the public to value the role of frontline workers in protecting people’s health (86). For example, local community members could send support and appreciation messages to healthcare workers (87). In addition, Healthcare workers should be fairly remunerated for their work. Mechanisms should be in place to ensure responders’ safety and well-being pre-, during, and post-deployment in an outbreak (86). In Table (3), we summarise the recommended actions derived from the existing literature and guidelines, that need to be in place to ensure the well-being of all responders involved in an outbreak response.

Our review identified a lack of RCCE interventions targeting vulnerable groups in the context of outbreaks. This finding is consistent with that of Bailey *et al.* (2023), who analysed 141 COVID-19 risk communication messages and found that less than 9 % of those messages are directed to vulnerable groups (i.e., older persons, people with psychosocial or physical disabilities), and less than 3 % of the messages addressed mental health. In addition, only two of the 26 RCTs identified in this review addressed children and adolescents, and none of the included RCTs addressed the caregivers’ MHPSS needs.

Several studies identified communication inequalities during outbreaks which led to considerable disparities in exposure to risk communication messages and hence in the adoption of IPC measures and the impact of the outbreak on different population groups (89–92). Communication inequality was associated with several factors such as age, income, education, ethnicity, disability, and language (89–92). Therefore, we recommend that RCCE interventions should consider these factors and be tailored to the needs of different population groups.

Inappropriate risk communication messages can trigger fear and anxiety among community members (93). During the Ebola outbreak in West Africa, risk communication messages focused on the lethality of the virus (such as “Ebola kills”) triggered fear, anxiety and hopelessness among affected community members. In contrast, messages focused on the importance of early treatment and survival improved health-seeking behaviours (93). Therefore, community representatives and different population groups should be involved in developing and delivering RCCE messages to ensure their appropriateness and acceptability.

Per the IASC recommendations, MHPSS activities implemented as a part of outbreak response should be adapted to minimise the risk of infection and ensure the continuity of services (17). In this review, we have presented examples from the literature where some adaptations and restrictions were made to ensure the safety of patients and providers (78–80). Although the use of technology to provide MHPSS interventions was found to be effective as it facilitates access to service, there is a concern that heavy reliance on technology may increase the disparity in health services, especially for people who are socially and digitally disadvantaged (30). Accordingly, this issue should be considered while developing and delivering MHPSS interventions.

In some outbreaks, such as EVD, MHPSS has a role in ensuring that burial is carried out in a safe and dignified manner. The IASC guidance for Ebola stresses that people should be allowed to mourn and practice their rituals without compromising IPC measures (e.g., from a safe distance without touching the body) (94). As a part of the International Federation of Red Cross and Red Crescent Societies (IFRC)’s response to the Ebola outbreak, staff trained in PFA provide condolence during burials and home visits to the families of deceased patients. This helps to gain trust and access to local communities (95).

Outbreaks challenge healthcare service efforts to provide care for infected individuals and simultaneously maintain services for other conditions. An assessment conducted by the WHO during COVID-19 found 93 % of countries reported disruption of mental health services due to the pandemic partially due to the diversion of the health system’s resources to COVID-19 (13). In many cases, mental health professionals were reallocated to work in Intensive Care Units and COVID-19 wards (96). In addition, several studies reported the suspension of mental health services to control and prevent infection (78–80). Such decisions will likely affect people with severe or acute mental health conditions who might need hospital admission. This indicates the importance of integrating MHPSS into general health services and exploring innovative ways to deliver MHPSS services. Despite their negative impact on mental health, emergencies such as outbreaks also present an opportunity to ‘Build Back Better’ mental health systems (97). Therefore, the role of MHPSS should not be limited to the response phase of public health emergencies but should also include preparedness and long-term recovery.

Despite the long-term psychosocial impacts of infection, most reviews focused on the acute response to infectious disease outbreaks, and none of the included interventions addressed long-term impacts. Therefore, attention should be given to those suffering from the long-term health effects of the infection. Active monitoring for psychological symptoms and referral to mental health services, when needed, were among the recommendations for post-COVID-19 rehabilitation (98).

Outbreaks and pandemics perpetuate the long-standing inequalities in healthcare among low-income communities and vulnerable groups (e.g., minority ethnic groups, children, refugees, migrants, and people with physical or psychosocial disabilities). This is evident when comparing COVID-19 deaths among minority ethnic groups. For instance, Black Americans accounted for 34% of confirmed COVID-19 cases, despite only comprising 13% of the US population (99). In the UK, the Bangladeshi group had the highest COVID-19 mortality rates during the second wave, which was 4-5 times higher than the White British group (100). Seemingly, children and young people were among the most affected by the COVID-19 pandemic, given the closure of schools, limited physical activity, lockdown, loss of loved ones due to infection, limited understanding of the situation, and spending hours online, which might have an impact on their mental health (32). Overall, there was a lack of MHPSS interventions targeting marginalised and vulnerable groups. Therefore, community-based MHPSS interventions should be developed to target marginalised and vulnerable groups, especially those who might not have access to health services.

Table (4) summarises research gaps we identified and the recommendations to narrow these gaps.

### Integrating MHPSS into outbreak response: a proposed framework

Given the immense impact of the COVID-19 pandemic, the WHO has stressed the importance of MHPSS “as an integral component in public health emergency response that must be addressed across a range of response pillars, including case management, risk communication and community engagement, and the maintenance of safe and accessible essential health services” (69). The WHO’s Incident Management System (IMS) is a standardised, yet flexible structure to managing a public health emergency response (68). As the IMS is the recognised structure for managing public health emergencies, including outbreaks, we used it to inform our proposed framework for integrating MHPSS into outbreak response (Figure 3). The framework summarises MHPSS operational considerations in relation to relevant IMS functions/sub-functions and outbreak response pillars.

**Figure 3:**
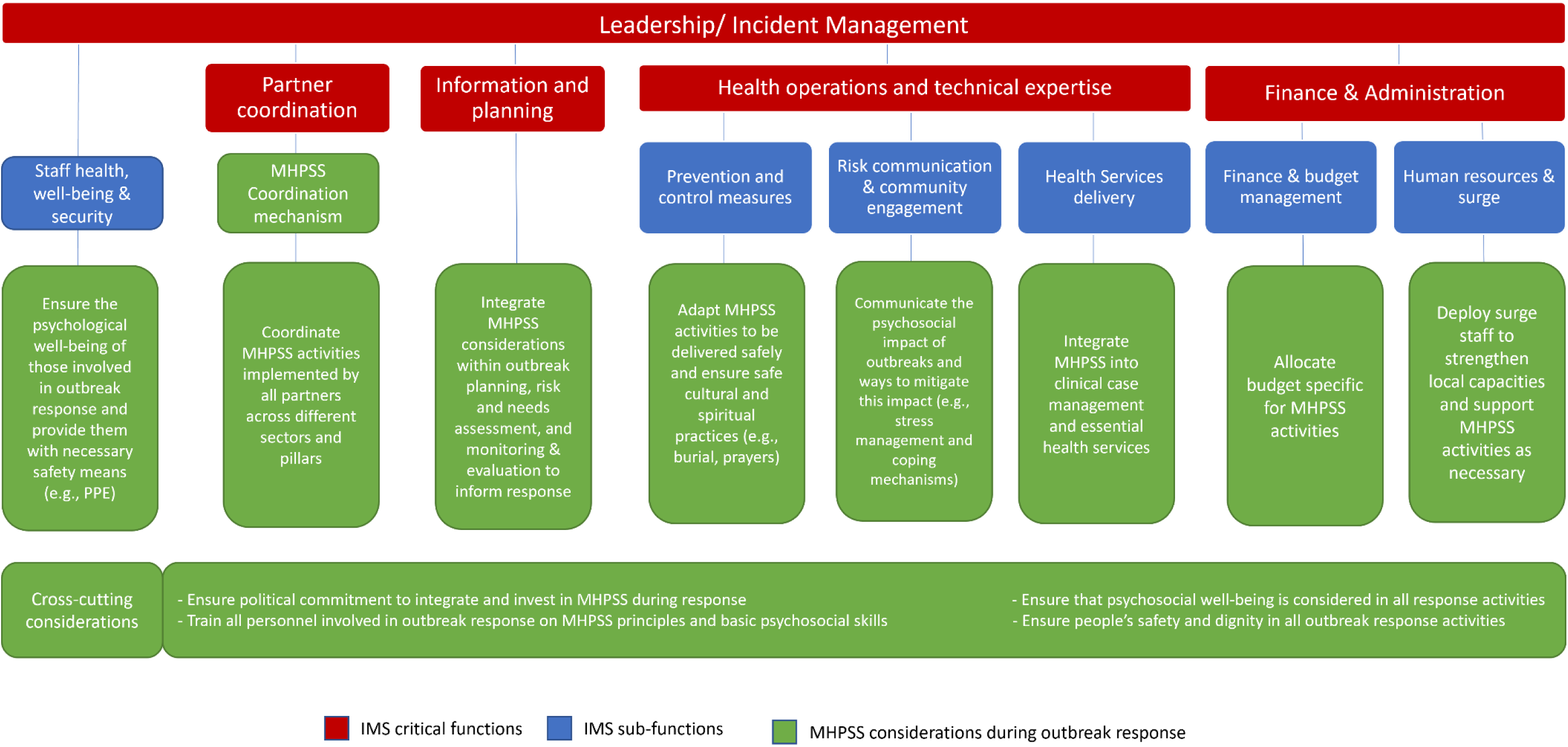
Operational framework to integrate MHPSS considerations into WHO’s Incident Management System (IMS)

Given that outbreak response is usually multi-sectoral, coordination becomes a necessity. For a better coordination between different actors involved in outbreak response, the IASC guidelines recommend establishing an intersectoral coordination group for MHPSS to ensure that psychological well-being is considered in all response activities (18). This becomes more evident during outbreaks, where many control measures can affect psychosocial well-being (e.g. quarantine and lockdown). While coordination can happen more readily between MHPSS actors, MHPSS actors need to work harder with non-MHPSS actors to ensure they integrate MHPSS considerations into their work. In Table 5, we recommend a set of actions to improve the MHPSS component of outbreaks preparedness and response, and hence mitigate their psychosocial impact.

**Table 3:** Recommended actions to ensure the well-being of responders before, during, and after deployment in an outbreak.

| Deployment phase | Recommended actions |
| --- | --- |
| Pre-deployment | • Screening and assessing the capacity of staff to respond to the emergency.<br>• Selecting the right persons with the right set of skills required for the posting.<br>• Capacity-strengthening including basic psychosocial skills and stress management/self-care skills besides technical skills required for the job.<br>• Risk communication including providing responders with up-to-date information about the outbreak, infectious agent, protection measures, available support mechanisms, and other information relevant to their safety and well-being. |
| During deployment | • Monitoring the outbreak's impact on responders' physical and mental health and managing these impacts, including through psychological support.<br>• Buddy system to provide both psychosocial and technical support for responders.<br>• Providing PPE and other tools needed to perform the job safely and appropriately.<br>• Ensuring that support is in place when needed and that responders are aware of these mechanisms (e.g., 24/7 helplines) and referral pathways to clinical services if required. |
| Post-deployment | • Post-deployment debriefing session for responders to share their experiences, feelings and thoughts and to assess if further support is needed.<br>• Monitoring possible delayed impacts of the stressful experience. |
| General considerations | • Valuing responders' efforts and reducing social stigma (e.g., appreciation and support from their organisations and local communities and remuneration for their work). |

**Table 4.** Identified research gaps and recommendations.

| Research gap | Recommendations |
| --- | --- |
| The majority of studies were conducted during the acute phase of outbreaks | Researchers should develop and evaluate interventions to address the long-term impact of outbreaks on well-being, as well as effectiveness of preparedness interventions |
| Most of the RCTs identified were conducted in healthcare settings targeting patients or healthcare workers. | Develop community-based interventions that target different population groups especially vulnerable groups |
| Research targeting frontline workers focused mainly on medical staff | Research targeting frontline workers should expand beyond healthcare to include others involved in outbreak response such as police and community volunteers |
| Technology was used heavily during COVID-19 to deliver MHPSS services, which might exclude disadvantaged population groups from accessing these services | Research needed to assess the acceptability and feasibility of those interventions and address the accessibility issues among vulnerable population groups |

**Table 5.** Recommendations for policy and practice.

| Recommendations | Examples from the literature |
| --- | --- |
| **General considerations** |  |
| Train all those involved in the outbreak response on PFA and basic psychosocial skills | In the Ebola outbreak in West Africa, health workers, community and faith leaders, police, contact tracers, volunteers, Ebola survivors, social mobilization agents and, food providers to quarantined households were trained in PFA (71) |
| Ensure that psychological well-being is considered in all outbreak response activities | Providing information and support during ambulance pick-ups, speaking to families of suspected and confirmed EVD patients, facilitating communication between families and patients, support to families of deceased patients, and ensure safe burials in a dignified manner (81) |
| **Coordination** |  |
| Establish clear multisectoral coordination mechanism to coordinate MHPSS across all response pillars and facilitate collaboration between different stakeholders. Link with IASC MHPSS Technical Working Group where established | Police forces were trained on PFA and nonviolent de-escalation techniques for agitated patients, especially those in Ebola Treatment Units (ETUs), and improved engagement with the public. The coordination between police and mental health staff facilitated referrals to mental health services for those needing further intervention (70). |
| **Staff/responders' well-being** |  |
| Establish support mechanisms for staff involved in outbreak response. | Peer support mechanisms, such as the "buddy system", can provide both psychosocial and technical support (72). |
| **Case management** |  |
| Ensure that MHPSS considerations are included in the clinical management guidelines for infected cases. | The guidance for clinical management of COVID-19 stated that "Basic psychosocial support skills are at the core of any clinical intervention for COVID-19. Such skills are indispensable for all involved in the COVID-19 clinical response, whether they identify as mental health and psychosocial providers or not." (101). |
| Address the long-term psychological impact of infection on the affected population. | Active monitoring for psychological symptoms and referral to mental health services, when needed, were among the recommendations for post-COVID-19 rehabilitation (98). |
| Train staff involved in clinical case management on basic psychosocial skills. | Those working in Ebola Treatment Units (ETUs), such as medical staff, Ebola survivors, police, and the Psychosocial support team received MHPSS training including PFA and self-care (70,71,81). |
| **Maintaining essential health services** |  |
| Ensure the integration of MHPSS into general health services to address the psychological needs of people with mental health and other chronic health conditions. | Nurses working in general hospitals were trained in PFA, case identification and referral pathways and provided basic counselling and problem-solving therapy for individuals needing mental healthcare (102). |
| **Community engagement and risk communications** |  |
| Communicate the psychosocial impact of outbreaks and ways to mitigate this impact (e.g., stress management). Adapt the messages to the different population groups' needs (e.g., children, people with disabilities, refugees). Involve community members in developing and delivering risk communication messages. | Text messages to support the population affected by the COVID-19 pandemic. Subscribers received free daily supportive messages for three months. Service users were involved in the development of these messages (83). |
| **Infection prevention and control** |  |
| Ensure that IPC measures are followed when delivering MHPSS interventions and adapt interventions to be delivered safely. | Adaptation of mental health services included postponing unnecessary outpatient visits and replacing them with check-ins via phones; restricting family visits; group activities were suspended or delivered remotely (e.g. using Zoom). Home hospitalisation model where people with severe mental illness who need admission were cared for at home by a mental health team (78–80). |
| Ensure safe and dignified burials and allow people to mourn and practice their rituals without compromising IPC measures. | Providing support to families of the deceased and ensuring safe and dignified burials (71) |

## Strengths and Limitations

To our knowledge, this is the first umbrella review to summarise review-level evidence. It is also the first review to address MHPSS interventions in relation to infectious disease outbreak response pillars. We believe that our findings and the proposed framework can guide the process of integrating MHPSS into future infectious disease outbreaks. However, our review has several limitations. The majority of the reviews included were of low quality, and the majority of included primary evidence included in those reviews was also of low quality. To mitigate the impact of this limitation, we focused on the evidence obtained from RCTs. (as reported in the included review). We did not carry out a quality appraisal for primary studies, but we included their quality as reported in the included reviews. Heterogeneity regarding study designs, interventions, and outcome measures was a major limitation for most of the included studies. The main causes of bias in the primary studies include lack of randomisation and blinding, small sample sizes, attrition bias, and selective reporting.

## Conclusion

Outbreaks have a substantial psychological impact on individuals, communities, and frontline workers. Therefore MHPSS should be an essential component of the outbreak response. Despite the low quality of the majority of the existing evidence, MHPSS interventions have the potential to improve the psychosocial well-being of those affected by and those responding to outbreaks. They also can improve the outcomes of the outbreak response activities such as contact tracing, infection prevention and control, and clinical case management. We proposed a framework for integrating MHPSS into outbreak response informed by the WHO’s IMS. The framework addresses MHPSS operational considerations that can help mitigate the mental health impact of outbreaks. Further research is needed to deepen the evidence for MHPSS interventions in infectious disease outbreaks.

## Data Availability

Not applicable

## Acknowledgements

We would like to thank members of the Africa public mental health consortium; Dr Florence Baingana (WHO AFRO), Dr. Mohammed Abdulaziz, Dr Adelard Kakunz, and Dumsani Njobo Mamba (Africa CDC) , and Rosemary Mwaisaka (ECSA-HC) for providing technical support throughout the project.

This review was conducted as a part of a project funded by the UK Public Health Rapid Support Team (UK-PHRST). The UK-PHRST is funded by UK Aid from the Department of Health and Social Care and is jointly run by UK Health Security Agency and the London School of Hygiene & Tropical Medicine. The views expressed in this publication are those of the author(s) and not necessarily those of the Department of Health and Social Care.

## Supplemental Files

S1: Detailed search strategy

S2: Detailed Quality appraisal results

