## Supplementary material for "Integrating Mental Health and Psycho-Social Support (MHPSS) into infectious disease outbreak and epidemic response: an umbrella review and operational framework": S2

| | Quality appraisal of the systematic revIew - JBI 2020 checklist | | | | | | | | | | | | |
|---|---|---|---|---|---|---|---|---|---|---|---|---|---|
| | 1. Is the review question clearly and explicitly stated? | 2. Were the inclusion criteria appropriate for the review | 3. Was the search strategy appropriate? | 4. Were the sources and resources used to search for studies adequate? | 5. Were the criteria for appraising studies appropriate? | 6. Was critical appraisal conducted by two or more reviewers | 7. Were there methods to minimize errors in data extraction? | 8. Were the methods used to combine studies appropriate? | 9. Was the likelihood of publication bias assessed? | 10. Were recommendations for policy and/or practice | 11. Were the specific directives for new research appropriate? | Total score | **Total Quality** |
| **Author, Year** | | | | | | | | | | | | | |
| **(Bertuzzi et al., 2021)** | 1 | 1 | 1 | 1 | 1 | 1 | 1 | 0 | 0 | 1 | 1 | 9 | High quality |
| **(Bursky et al., 2021)** | 1 | 1 | 0 | 1 | 1 | 1 | 0 | 0 | 0 | 1 | 1 | 7 | Moderate quality |
| **(Cénat, Felix, et al., 2020)** | 1 | 1 | 1 | 1 | 0 | 0 | 0 | 0 | 0 | 1 | 0 | 5 | Low quality |
| **(Damiano et al., 2021)** | 1 | 0 | 1 | 1 | 1 | 1 | 0 | 1 | 0 | 1 | 0 | 7 | Moderate quality |
| **(Davison et al., 2021)** | 1 | 1 | 1 | 1 | 1 | 0 | 0 | 0 | 0 | 1 | 1 | 7 | Moderate quality |
| **(Ding et al., 2021)** | 1 | 1 | 1 | 1 | 1 | 1 | 0 | 1 | 1 | 0 | 1 | 9 | High quality |
| **(Doherty et al., 2021)** | 1 | 1 | 1 | 1 | 1 | 1 | 1 | 1 | 0 | 1 | 1 | 10 | High quality |
| **(Gómez et al., 2021)** | 1 | 1 | 1 | 0 | 0 | 0 | 0 | 0 | 0 | 1 | 0 | 4 | Low quality |
| **(Hooper et al., 2021)** | 1 | 1 | 1 | 1 | 0 | 0 | 0 | 0 | 0 | 1 | 0 | 5 | low quality |
| **(Kunzler et al., 2021)** | 1 | 1 | 1 | 1 | 0 | 0 | 1 | 0 | 0 | 1 | 1 | 7 | Moderate quality |
| **(Meherali et al., 2021)** | 1 | 1 | 1 | 1 | 1 | 0 | 1 | 0 | 0 | 1 | 1 | 8 | Moderate quality |
| **(Pollock et al., 2020)** | 1 | 1 | 1 | 1 | 1 | 1 | 1 | 0 | 0 | 1 | 1 | 9 | High quality |
| **(Puyat et al., 2020)** | 1 | 1 | 1 | 1 | 1 | 0 | 0 | 0 | 0 | 1 | 1 | 7 | Moderate quality |
| **(Rivera-Torres et al., 2021)** | 1 | 0 | 1 | 1 | 0 | 0 | 0 | 0 | 0 | 0 | 0 | 3 | Low quality |
| **(Serrano-Ripoll et al., 2020)** | 1 | 1 | 1 | 1 | 1 | 0 | 1 | 1 | 1 | 1 | 1 | 10 | High quality |
| **(Shatri et al., 2021)** | 0 | 1 | 1 | 1 | 1 | 1 | 0 | 0 | 0 | 0 | 0 | 5 | Low quality |
| **(Soklaridis et al., 2020)** | 1 | 1 | 1 | 1 | 1 | 1 | 0 | 0 | 0 | 0 | 1 | 7 | Moderate quality |
| **(Strudwick et al., 2021)** | 1 | 1 | 1 | 1 | 0 | 0 | 0 | 0 | 0 | 1 | 1 | 6 | Moderate quality |
| **(Sun et al., 2021)** | 1 | 1 | 1 | 1 | 0 | 1 | 0 | 1 | 0 | 0 | 1 | 7 | Moderate quality |
| **(Williams et al., 2021)** | 1 | 1 | 1 | 1 | 1 | 1 | 0 | 0 | 0 | 1 | 1 | 8 | Moderate quality |
| **(Yang et al., 2021)** | 1 | 1 | 1 | 1 | 1 | 1 | 1 | 1 | 0 | 0 | 0 | 8 | Moderate quality |
| **(Yue et al., 2020)** | 1 | 1 | 1 | 1 | 1 | 1 | 0 | 0 | 0 | 1 | 1 | 8 | Moderate quality |
| **(Zaçe et al., 2021)** | 1 | 0 | 1 | 1 | 0 | 1 | 0 | 0 | 0 | 1 | 1 | 6 | Moderate quality |
