## Supplementary material for "Integrating Mental Health and Psycho-Social Support (MHPSS) into infectious disease outbreak and epidemic response: an umbrella review and operational framework": S1

**Detailed Search Strategy**

**SCOPUS**

( TITLE-ABS-KEY ( ( "Psychosocial Intervention*" OR "Psycho-social Intervention*" OR "Psychological Intervention*" OR Psychotherapy OR Psychotherapies OR "Psychiatric Intervention*" OR "Psychological Treatment*" OR "Mental Health Service*" OR "Mental Health Intervention*" ) ) AND TITLE-ABS-KEY ( ( "Disease Outbreak" OR Outbreaks OR "Infectious Disease Outbreak*" OR Pandemic* OR Epidemic* OR Quarantine OR SARS OR "Severe Acute Respiratory Syndrome" OR "SARS-CoV" OR "Middle East Respiratory Syndrome Coronavirus" OR "myers-cov" OR myers OR COVID-19 OR "SARS-CoV-2" OR "Ebola Virus" OR "Ebola Hemorrhagic Fever" OR "inluenza a h1n1" OR "hand virus" ) ) AND TITLE-ABS-KEY ( ( Stress OR Depression OR Anxiety OR "Post-Traumatic Stress Disorders" OR PTSD OR "Psychological Resilience" OR "Psychological Distress" OR Coping OR "Psychological Adaptation" OR "Adjustment" OR "Coping Skills" OR "Coping Strategies" OR "Psychological Wellbeing" OR "Psychological Well-being" ) ) AND TITLE-ABS-KEY ( ( "systematic review" OR "systematic literature review" OR "scoping review" OR "systematic narrative review" OR "systematic qualitative review" OR "systematic evidence review" OR "systematic quantitative review" OR "systematic meta-review" OR "systematic critical review" OR "systematic mixed studies review" OR "systematic mapping review" OR "systematic Cochrane review" OR "systematic search and review" OR "systematic integrative review" OR "rapid review" OR "Rapid systematic review" OR meta-analysis ) ) )

**Scopus search results =135**

**PubMed**

((("Psychosocial Intervention*" OR "Psycho-social Intervention*" OR "Psychological Intervention*" OR Psychotherapy OR Psychotherapies OR "Psychiatric Intervention*" OR "Psychological Treatment*" OR "Mental Health Service*" OR "Mental Health Intervention*") AND ("Disease Outbreak" OR Outbreaks OR "Infectious Disease Outbreak*" OR Pandemic* OR Epidemic* OR Quarantine OR SARS OR "Severe Acute Respiratory Syndrome" OR "SARS-CoV" OR "Middle East Respiratory Syndrome Coronavirus" OR "MERS-CoV" OR MERS OR COVID-19 OR "SARS-CoV-2" OR "Ebola Virus" OR "Ebola Hemorrhagic Fever" OR "Inﬂuenza A H1N1" OR "H1N1 Virus") ) AND (Stress OR Depression OR Depressive OR Anxiety OR "Post-Traumatic Stress Disorders" OR PTSD OR "Psychological Resilience" OR "Psychological Distress" OR Coping OR "Psychological Adaptation" OR "Adjustment" OR "Coping Skills" OR "Coping Strategies" OR "Psychological Wellbeing" OR "Psychological Well-being") ) AND ("systematic review"[Title/Abstract] OR "systematic literature review"[Title/Abstract] OR "scoping review"[Title/Abstract] OR "systematic narrative review"[Title/Abstract] OR "systematic qualitative review"[Title/Abstract] OR "systematic evidence review"[Title/Abstract] OR "systematic quantitative review"[Title/Abstract] OR "systematic meta-review"[Title/Abstract] OR "systematic critical review"[Title/Abstract] OR "systematic mixed studies review"[Title/Abstract] OR "systematic mapping review"[Title/Abstract] OR "systematic Cochrane review"[Title/Abstract] OR "systematic search and review"[Title/Abstract] OR "systematic integrative review"[Title/Abstract] OR "rapid review"[Title/Abstract] OR "Rapid systematic review"[Title/Abstract] OR meta-analysis[Title/Abstract])

**PubMed search results = 757**

**WOS**

("Psychosocial Intervention*" OR "Psycho-social Intervention*" OR "Psychological Intervention*" OR Psychotherapy OR Psychotherapies OR "Psychiatric Intervention*" OR "Psychological Treatment*" OR "Mental Health Service*" OR "Mental Health Intervention*") (All Fields) and (Stress OR Depression OR Anxiety OR "Post-Traumatic Stress Disorder*" OR PTSD OR "Psychological Resilience" OR "Psychological Distress" OR Coping OR "Psychological Adaptation" OR "Adjustment" OR "Coping Skills" OR "Coping Strategies" OR "Psychological wellbeing” OR “Psychological well-being”) (All Fields) and ("Disease Outbreak" OR Outbreaks OR "Infectious Disease Outbreak*" OR Pandemic* OR Epidemic* OR Quarantine OR SARS OR "Severe Acute Respiratory Syndrome" OR "SARS-CoV" OR "Middle East Respiratory Syndrome Coronavirus" OR "MERS-CoV" OR MERS OR COVID-19 OR "SARS-CoV-2" OR "Ebola Virus" OR "Ebola Hemorrhagic Fever" OR "Inﬂuenza A H1N1" OR "H1N1 Virus") (All Fields) and ("systematic review" OR "systematic literature review" OR "scoping review" OR "systematic narrative review" OR "systematic qualitative review" OR "systematic evidence review" OR "systematic quantitative review" OR "systematic meta-review" OR "systematic critical review" OR "systematic mixed studies review" OR "systematic mapping review" OR "systematic Cochrane review" OR "systematic search and review" OR "systematic integrative review" OR "rapid review" OR "Rapid systematic review" OR meta-analysis) (All Fields)

**WOS results = 98**

Ovid MEDLINE(R) ALL <1946 to December 30, 2021>

1	(psychosocial or psycho-social or psychological or "mental health" or wellbeing or well-being or mental illness* or mental disorder*).mp. [mp=title, abstract, original title, name of substance word, subject heading word, floating sub-heading word, keyword heading word, organism supplementary concept word, protocol supplementary concept word, rare disease supplementary concept word, unique identifier, synonyms]	1070385

2	(Stress or Depression or Depressive or Anxiety or "Post-Traumatic Stress Disorders" or PTSD or "Psychological Resilience" or "Psychological Distress" or Coping or "Psychological Adaptation" or "Adjustment" or "Coping Skills" or "Coping Strategies").mp. [mp=title, abstract, original title, name of substance word, subject heading word, floating sub-heading word, keyword heading word, organism supplementary concept word, protocol supplementary concept word, rare disease supplementary concept word, unique identifier, synonyms]	1798575

3	mental health/ or exp mental disorders/	1375047

4	1 or 2 or 3	3232569

5	("Psychosocial Intervention*" or "Psycho-social Intervention*" or "Psychological Intervention*" or Psychotherapy or Psychotherapies or "Psychiatric Intervention*").mp. [mp=title, abstract, original title, name of substance word, subject heading word, floating sub-heading word, keyword heading word, organism supplementary concept word, protocol supplementary concept word, rare disease supplementary concept word, unique identifier, synonyms]	101215

6	exp mental health services/ or community mental health services/ or exp counseling/ or emergency services, psychiatric/ or exp psychotherapy/	301693

7	5 or 6	320715

8	4 and 7	218116

9	("Disease Outbreak" or Outbreaks or "Infectious Disease Outbreak*" or Pandemic* or Epidemic* or Quarantine or SARS or "Severe Acute Respiratory Syndrome" or "SARS-CoV" or "Middle East Respiratory Syndrome Coronavirus" or "MERS-CoV" or MERS or COVID-19 or "SARS-CoV-2" or "Ebola Virus" or "Ebola Hemorrhagic Fever" or "Inﬂuenza A H1N1" or "H1N1 Virus").mp. [mp=title, abstract, original title, name of substance word, subject heading word, floating sub-heading word, keyword heading word, organism supplementary concept word, protocol supplementary concept word, rare disease supplementary concept word, unique identifier, synonyms]	460745

10	disease outbreaks/ or disease hotspot/ or epidemics/ or pandemics/	169494

11	9 or 10	460777

12	4 and 7 and 11	2341

13	("systematic review" or "systematic literature review" or "scoping review" or "systematic narrative review" or "systematic qualitative review" or "systematic evidence review" or "systematic quantitative review" or "systematic meta-review" or "systematic critical review" or "systematic mixed studies review" or "systematic mapping review" or "systematic Cochrane review" or "systematic search and review" or "systematic integrative review" or "rapid review" or "Rapid systematic review").mp. [mp=title, abstract, original title, name of substance word, subject heading word, floating sub-heading word, keyword heading word, organism supplementary concept word, protocol supplementary concept word, rare disease supplementary concept word, unique identifier, synonyms]	254861

14	exp "Systematic Review"/	180624

15	exp meta-analysis/	149724

16	13 or 14 or 15	324975

17	12 and 16	106

Results : 106

Hit : 3/1/2022

**APA PsycINFO <1806 to December Week 4 2021>**

1	(psychosocial or psycho-social or psychological or "mental health" or wellbeing or well-being or mental illness* or mental disorder*).mp. [mp=title, abstract, heading word, table of contents, key concepts, original title, tests & measures, mesh word]	990945

2	(Stress or Depression or Depressive or Anxiety or "Post-Traumatic Stress Disorders" or PTSD or "Psychological Resilience" or "Psychological Distress" or Coping or "Psychological Adaptation" or "Adjustment" or "Coping Skills" or "Coping Strategies").mp. [mp=title, abstract, heading word, table of contents, key concepts, original title, tests & measures, mesh word]	858068

3	exp mental disorders/ or exp mental health/	970194

4	1 or 2 or 3	1920221

5	("Psychosocial Intervention*" or "Psycho-social Intervention*" or "Psychological Intervention*" or Psychotherapy or Psychotherapies or "Psychiatric Intervention*").mp. [mp=title, abstract, heading word, table of contents, key concepts, original title, tests & measures, mesh word]	170101

6	mental health/ or behavioral health services/ or mental health programs/ or mental health services/ or preventive mental health services/	112981

7	exp psychotherapy/	211816

8	exp counseling/	80324

9	intervention/ or crisis intervention/ or early intervention/ or family intervention/ or group intervention/ or school based intervention/ or workplace intervention/	116957

10	5 or 6 or 7 or 8	419525

11	("Disease Outbreak" or Outbreaks or "Infectious Disease Outbreak*" or Pandemic* or Epidemic* or Quarantine or SARS or "Severe Acute Respiratory Syndrome" or "SARS-CoV" or "Middle East Respiratory Syndrome Coronavirus" or "MERS-CoV" or MERS or COVID-19 or "SARS-CoV-2" or "Ebola Virus" or "Ebola Hemorrhagic Fever" or "Inﬂuenza A H1N1" or "H1N1 Virus").mp. [mp=title, abstract, heading word, table of contents, key concepts, original title, tests & measures, mesh word]	30427

12	exp disease outbreaks/	9324

13	exp epidemics/	9226

14	exp pandemics/	5835

15	11 or 12 or 13 or 14	30427

16	("systematic review" or "systematic literature review" or "scoping review" or "systematic narrative review" or "systematic qualitative review" or "systematic evidence review" or "systematic quantitative review" or "systematic meta-review" or "systematic critical review" or "systematic mixed studies review" or "systematic mapping review" or "systematic Cochrane review" or "systematic search and review" or "systematic integrative review" or "rapid review" or "Rapid systematic review").mp. [mp=title, abstract, heading word, table of contents, key concepts, original title, tests & measures, mesh word]	40724

17	exp "systematic review"/	672

18	exp Meta Analysis/	5125

19	exp "literature review"/	23580

20	16 or 17 or 18 or 19	67522

21	4 and 10 and 15 and 20	85

**APA PsycINFO Results: 85**

**Global Health <1910 to 2021 Week 50>**

1	(psychosocial or psycho-social or psychological or "mental health" or wellbeing or well-being or "mental illness*" or "mental disorder*").mp. [mp=abstract, title, original title, broad terms, heading words, identifiers, cabicodes]	138843

2	(Stress or Depression or Depressive or Anxiety or "Post-Traumatic Stress Disorders" or PTSD or "Psychological Resilience" or "Psychological Distress" or Coping or "Psychological Adaptation" or "Adjustment" or "Coping Skills" or "Coping Strategies").mp. [mp=abstract, title, original title, broad terms, heading words, identifiers, cabicodes]	227612

3	exp mental disorders/ or exp mental health/ or exp mental stress/	105604

4	1 or 2 or 3	328248

5	("Psychosocial Intervention*" or "Psycho-social Intervention*" or "Psychological Intervention*" or Psychotherapy or Psychotherapies or "Psychiatric Intervention*").mp. [mp=abstract, title, original title, broad terms, heading words, identifiers, cabicodes]	3473

6	exp psychotherapy/	3850

7	exp counselling/	12259

8	5 or 6 or 7	17729

9	exp outbreaks/	62962

10	exp pandemics/ or exp epidemics/	74661

11	("Disease Outbreak" or Outbreaks or "Infectious Disease Outbreak*" or Pandemic* or Epidemic* or Quarantine or SARS or "Severe Acute Respiratory Syndrome" or "SARS-CoV" or "Middle East Respiratory Syndrome Coronavirus" or "MERS-CoV" or MERS or COVID-19 or "SARS-CoV-2" or "Ebola Virus" or "Ebola Hemorrhagic Fever" or "Inﬂuenza A H1N1" or "H1N1 Virus").mp. [mp=abstract, title, original title, broad terms, heading words, identifiers, cabicodes]	210635

12	9 or 10 or 11	210635

13	4 and 8 and 12	671

14	("systematic review" or "systematic literature review" or "scoping review" or "systematic narrative review" or "systematic qualitative review" or "systematic evidence review" or "systematic quantitative review" or "systematic meta-review" or "systematic critical review" or "systematic mixed studies review" or "systematic mapping review" or "systematic Cochrane review" or "systematic search and review" or "systematic integrative review" or "rapid review" or "Rapid systematic review").mp. [mp=abstract, title, original title, broad terms, heading words, identifiers, cabicodes]	45540

15	exp systematic reviews/ or exp meta-analysis/	59840

16	14 or 15	67594

17	13 and 16	26

**Global Health Results: 26**

**Embase <1974 to 2021 December 30>**

1	(psychosocial or psycho-social or psychological or "mental health" or wellbeing or well-being or "mental illness*" or "mental disorder*").mp. [mp=title, abstract, heading word, drug trade name, original title, device manufacturer, drug manufacturer, device trade name, keyword heading word, floating subheading word, candidate term word]	1355750

2	(Stress or Depression or Depressive or Anxiety or "Post-Traumatic Stress Disorders" or PTSD or "Psychological Resilience" or "Psychological Distress" or Coping or "Psychological Adaptation" or "Adjustment" or "Coping Skills" or "Coping Strategies").mp. [mp=title, abstract, heading word, drug trade name, original title, device manufacturer, drug manufacturer, device trade name, keyword heading word, floating subheading word, candidate term word]	2511451

3	exp mental health/ or exp psychological well-being/	186903

4	1 or 2 or 3	3403818

5	("Psychosocial Intervention*" or "Psycho-social Intervention*" or "Psychological Intervention*" or Psychotherapy or Psychotherapies or "Psychiatric Intervention*").mp. [mp=title, abstract, heading word, drug trade name, original title, device manufacturer, drug manufacturer, device trade name, keyword heading word, floating subheading word, candidate term word]	122698

6	exp mental health service/ or exp community mental health service/	59716

7	exp psychiatric treatment/ or exp "psychological and psychiatric procedures"/	1316353

8	exp psychotherapy/	268257

9	exp counseling/	182895

10	5 or 6 or 7 or 8 or 9	1475717

11	4 and 10	904260

12	("Disease Outbreak" or Outbreaks or "Infectious Disease Outbreak*" or Pandemic* or Epidemic* or Quarantine or SARS or "Severe Acute Respiratory Syndrome" or "SARS-CoV" or "Middle East Respiratory Syndrome Coronavirus" or "MERS-CoV" or MERS or COVID-19 or "SARS-CoV-2" or "Ebola Virus" or "Ebola Hemorrhagic Fever" or "Inﬂuenza A H1N1" or "H1N1 Virus").mp. [mp=title, abstract, heading word, drug trade name, original title, device manufacturer, drug manufacturer, device trade name, keyword heading word, floating subheading word, candidate term word]	491633

13	exp epidemic/	117689

14	exp pandemic/	100157

15	12 or 13 or 14	491786

16	4 and 10 and 15	12766

17	("systematic review" or "systematic literature review" or "scoping review" or "systematic narrative review" or "systematic qualitative review" or "systematic evidence review" or "systematic quantitative review" or "systematic meta-review" or "systematic critical review" or "systematic mixed studies review" or "systematic mapping review" or "systematic Cochrane review" or "systematic search and review" or "systematic integrative review" or "rapid review" or "Rapid systematic review").mp. [mp=title, abstract, heading word, drug trade name, original title, device manufacturer, drug manufacturer, device trade name, keyword heading word, floating subheading word, candidate term word]	425596

18	exp "systematic review"/	325982

19	exp meta analysis/	233834

20	17 or 18 or 19	519306

21	16 and 20	503

**Embase Results: 503**
